# Hepatitis E in Kathmandu Valley: Insights from a Representative Longitudinal Serosurvey

**DOI:** 10.1101/2023.11.28.23299131

**Authors:** Nishan Katuwal, Melina Thapa, Sony Shrestha, Krista Vaidya, Isaac I Bogoch, Jason Andrews, Rajeev Shrestha, Dipesh Tamrakar, Kristen Aiemjoy

**Affiliations:** Research and Development Division, Dhulikhel Hospital, Kathmandu University Hospital, Nepal; Center for Infectious Disease Research and Surveillance, Dhulikhel Hospital, Kathmandu University Hospital, Nepal; Division of Epidemiology, Department of Public Health Sciences, University of California Davis School of Medicine, USA; Department of Medicine, University of Toronto, Canada; Division of Infectious Diseases and Geographic Medicine, Stanford University School of Medicine, USA; Department of Community Medicine, Kathmandu University School of Medical Sciences, Nepal; Department of Microbiology and Immunology, Mahidol University Faculty of Tropical Medicine, Thailand

**Author notes:** **Corresponding Author:** Dr. Kristen Aiemjoy. equal contribution (Dipesh Tamrakar, Kristen Aiemjoy).

## Abstract

Hepatitis-E virus (HEV), an etiologic agent of acute inflammatory liver disease, is a significant cause of morbidity and mortality in South Asia. HEV is considered endemic in Nepal; but data on population-level infection transmission is sparse. We conducted a representative longitudinal serologic study between February 2019 and April 2021 in urban and peri-urban areas of central Nepal to characterize community-level HEV transmission. Individuals were followed up to four times, during which capillary blood samples were collected on dried blood spots and tested for anti-HEV immunoglobulin-G antibodies. Analyzing 2513 dried blood samples from 923 participants aged 0-25 years, we found a seroprevalence of 4.8% and a seroincidence rate of 10.9 per 1000 person-years. Notably, young adults, including women of childbearing age, faced the highest incidence of infection. Geospatial analysis identified potential HEV clusters in Kavre and Kathmandu districts, emphasizing the need for targeted interventions. Water source played a crucial role in HEV transmission, with individuals consuming surface water facing the highest risk of seroconversion. Our findings underscore the endemic nature of HEV in Nepal, emphasizing the importance of safe water practices and potential vaccination strategies for high-risk groups.

## BACKGROUND

Hepatitis E Virus (HEV), first identified in 1983, has emerged as the leading cause of acute clinical hepatitis in South Asia (1). The overall mortality rate associated with HEV is 1–4% (2). Immunocompromised individuals and pregnant women face the highest risk for complications and death, with mortality rates reaching up to 20% among pregnant women in their third trimester (3). The virus is primarily transmitted through food and water contaminated with infected fecal material, disproportionately affecting individuals in locations lacking improved sanitation systems (2).

HEV is estimated to cause approximately 20 million new infections annually, leading to around 3.3 million symptomatic cases and 70,000 deaths (4). However, these figures may be underestimates due to limited surveillance capacity and suboptimal access to laboratory diagnostics, which often leave many infections, especially pauci-symptomatic and subclinical cases, undetected and under-reported (5).

Sero-epidemiology provides a valuable tool to augment clinical surveillance of HEV, particularly when diagnostics and reporting systems are limited (6). Population-level sero-responses also offers insights into exposure that are not biased by access to health care and care-seeking behaviors (7). Despite the existence of four recognized HEV genotypes, the antibody responses they elicit are similar (2). IgG antibody responses peak 2–6 weeks post-infection (8,9) then start to decay. A study of confirmed HEV patients in Nepal found that IgG responses decayed substantially in the first 6 months but remained elevated above a seropositivity threshold for at least 14 months (10).

Nepal, considered endemic for hepatitis E virus, has experienced repeated outbreaks of HEV with sporadic cases of acute hepatitis between such outbreaks (11). A study among individuals seeking orthopedic care in Kathmandu between 2010 and 2012 found an IgG seroprevalence of 47.1% (12). Among healthy blood donors in Kathmandu in 2014, the age-adjusted seroprevalence of HEV was 3.2% for IgM and 8.3% for IgG (13). A study conducted after the 2015 earthquake among blood donors residing in earthquake-affected areas found a HEV seroprevalence of 3.2% for IgM and 41.9% for IgG (14). Most previous seroprevalence studies were conducted using convenience samples (healthy blood donors or individuals seeking care in a hospital setting), and it is unknown how well these estimates represent the general population.

To meet this gap, we performed a longitudinal HEV serosurvey among a geographically random population-based sample of children and young adults residing in Kathmandu and Kavre districts. The aim was to gain a better understanding of the geographic distribution of HEV, characterize the incidence, and investigate risk factors related to exposure.

## METHODS

### Overview

We conducted a representative, population-based longitudinal cohort study in urban (Kathmandu) and peri-urban (Kavre) areas of Nepal to characterize community-level HEV incidence. We enrolled geographically random sample of individuals aged 0-25 years from the catchment areas of Kathmandu Medical College in Kathmandu and Dhulikhel Hospital in Kavrepalanchok, Nepal from Feb 2019 to Apr 2021(15). Participants were followed-up three times, approximately 3 months, 6 months, and 12 months after initial enrollment and consent. During enrollment, relevant demographic data including age, sex, socioeconomic status, were collected.

### Sampling method

We employed a systematic random sampling strategy within our defined catchment areas. We randomly selected grid clusters, enumerated all households within each cluster, and then randomly selected participants based on age stratification using the groups 0-4, 5-9, 10-15, and 16-25 years. Our exclusion criteria were minimal to ensure a representative sample. We only excluded individuals who were not residents of the catchment area or did not fall within our specified age range at the time of the cross-sectional survey.

### Sample collection

At each visit, we collected capillary blood samples onto filter papers (TropBio FP 05-002-12) using a finger-prick (dried blood spots; DBS). The samples were dried at room temperature for 24 hours then stored in individual plastic bags with desiccant at −20 C.

### Laboratory Methods

We eluted the dried blood spots by submerging a single filled filter paper protrusion in 133 µL of 1X PBS containing 0.05% Tween buffer and incubating overnight at 4°C. We then centrifuged the tubes at 10,000xg and aliquoted the supernatant, considering this DBS eluate as a 1:10 dilution.

We analyzed the samples using end-point ELISA with recomWell HEV IgG ELISA kits (Mikrogen Diagnostik), which evaluate immunoglobulin IgG against the recombinant HEV-ORF2 antigen. We read the ELISA plates at a wavelength of 450nm (with a reference wavelength of 690nm) using a Bio-Rad iMark ELISA plate reader.

### Statistical Analysis

We fit 2-component Gaussian finite mixture models to determine an optimal cutoff value, defined as the mean plus 3.5 standard deviations from the lower mixture component of the log-transformed values (16). We then calculated seroprevalence as the percentage of samples that were seropositive at each sampling interval. To describe how seropositivity changes over age, we fit generalized additive models with a cubic spline for age (17). We computed simultaneous confidence intervals using a parametric bootstrap of the variance-covariance matrix of the fitted model parameters (18) (Mara, 2012). To investigate variables associated with seropositivity, we used mixed-effect binomial logit models with a random effect for community (19).

We calculated seroincidence, defined as the number of new seroconversions per 1000 person years, by dividing the number of individuals who seroconverted by the person-time at risk between measurements. We defined incident seroconversions as a change in IgG across the seropositivity cutoff between sampling intervals. We used negative binomial mixed-effects regression models to explore the influence of various predictors on seroincidence, including area, gender, income level, water source, and frequency of eating outside, while adjusting for age. Each predictor was individually incorporated into the model, which also accounted for repeated measurements on the same individual across different visits and individual-level variability. An offset was introduced to adjust for person-time.

### Ethical Considerations

The study protocol was reviewed and approved by the Nepal Health Research Council and Ethics Review Board of Dhulikhel Hospital and Stanford University. Written informed consent was obtained from all participants or their parents or legal guardians in the case of minors age <18. In addition, we obtained written assent from children between the ages of 15 and 17. The study was conducted following the principles of the Declaration of Helsinki.

## RESULTS

We collected a total of 2513 dried blood samples from 923 study participants between February 2019 to April 2021. Out of 923 study participants, 401 were from Kathmandu, 197 from Panauti (Kavre), 167 from Banepa (Kavre), 85 from Dhulikhel (Kavre) and 73 from Panchkhal (Kavre). The median age of participants was 11 years (Inter Quartile Range [IQR]: 5.6 - 17) and 52.2% (482/923) were male. The median number of study visits completed was 3 (IQR 2-3); 229/923; 65% (603/923) of participants had 3 or more study visits while 18.2% 168/923 had just 1 study visit and 16.4% 152/923 had 2 study visits (Table 1).

**Table 1:**
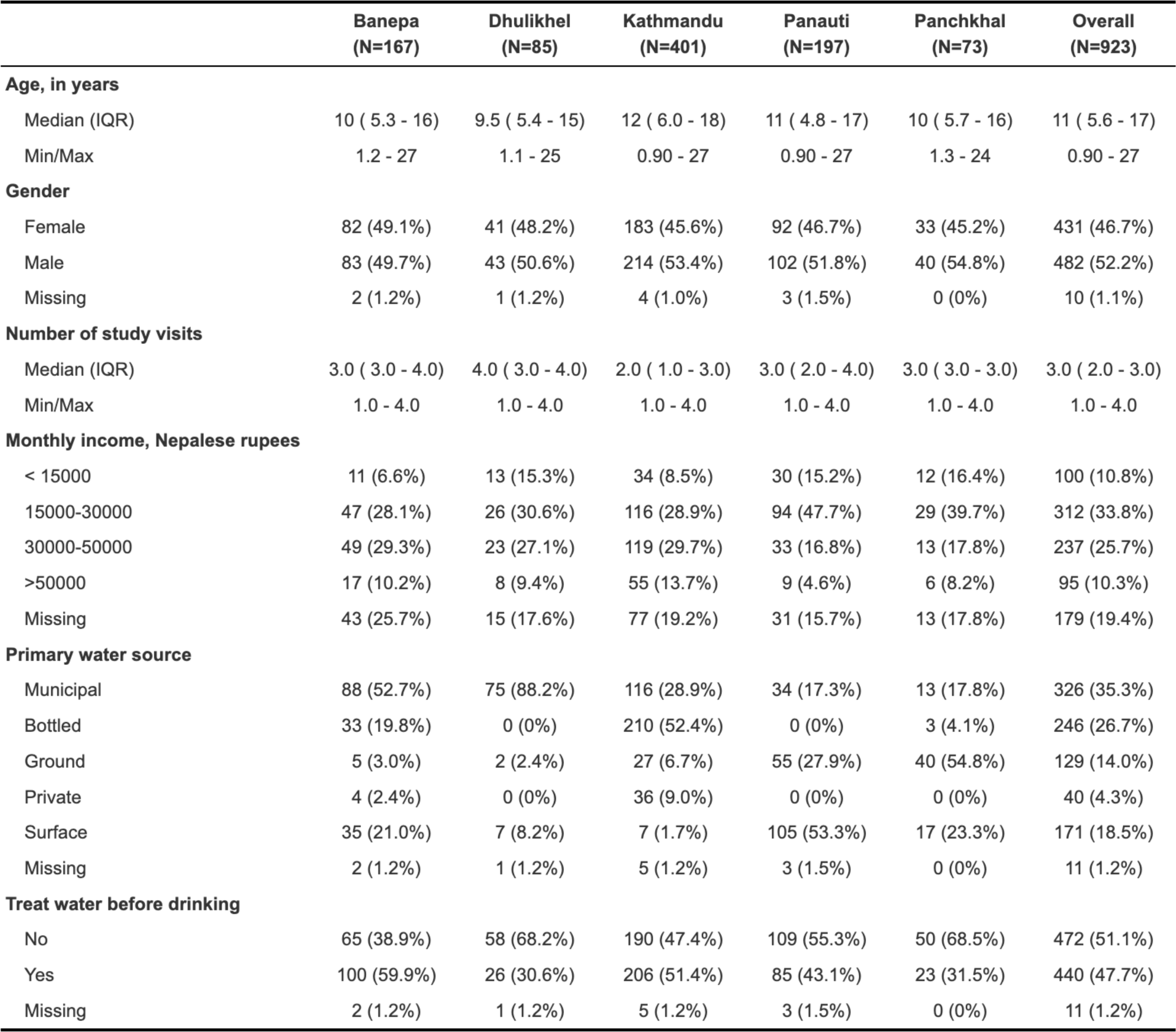
Demographic Characteristics of enrolled individuals.

From the total of 2513 processed samples from 923 enrolled individuals, 106 samples (53 individuals) were seropositive for HEV (Figure 1). At the baseline visit, the crude seroprevalence was 4.8% (44/923) (Table 2). There were 9 incident seroconversions over 822.8 years of person-time, yielding a crude seroincidence rate of 10.9 (95% CI 5-20.8) per 1000 person-years (Table 3). Both seroprevalence and seroincidence increased with age. Among 0-5 year olds, the seroprevalence was 1.0% (2/199), increasing to 11.1% (33/297) in the 15 to 25-year-old age group (p = 0.001; see Table 2 and Figure 2). The seroincidence rate rose from 0 (95%CI: 0 - 32) per 1000 person-years for 0 to 5 year-olds to 9.0 (95%CI: 1 - 32.4) for 5 to 10 year-olds, 14.0 (95%CI: 2.9 - 40.8) for 10 to 15 year-olds, and 14.8 (95%CI: 4.0 - 37.9) for 15 to 25 year-olds (Table 3). The age-adjusted seroincidence was similar among females (10.3, 95% CI 2.8 - 26.3) compared to males (11.6, 95% CI 3.8 - 27.2) (Table 3).

**Figure 1:**
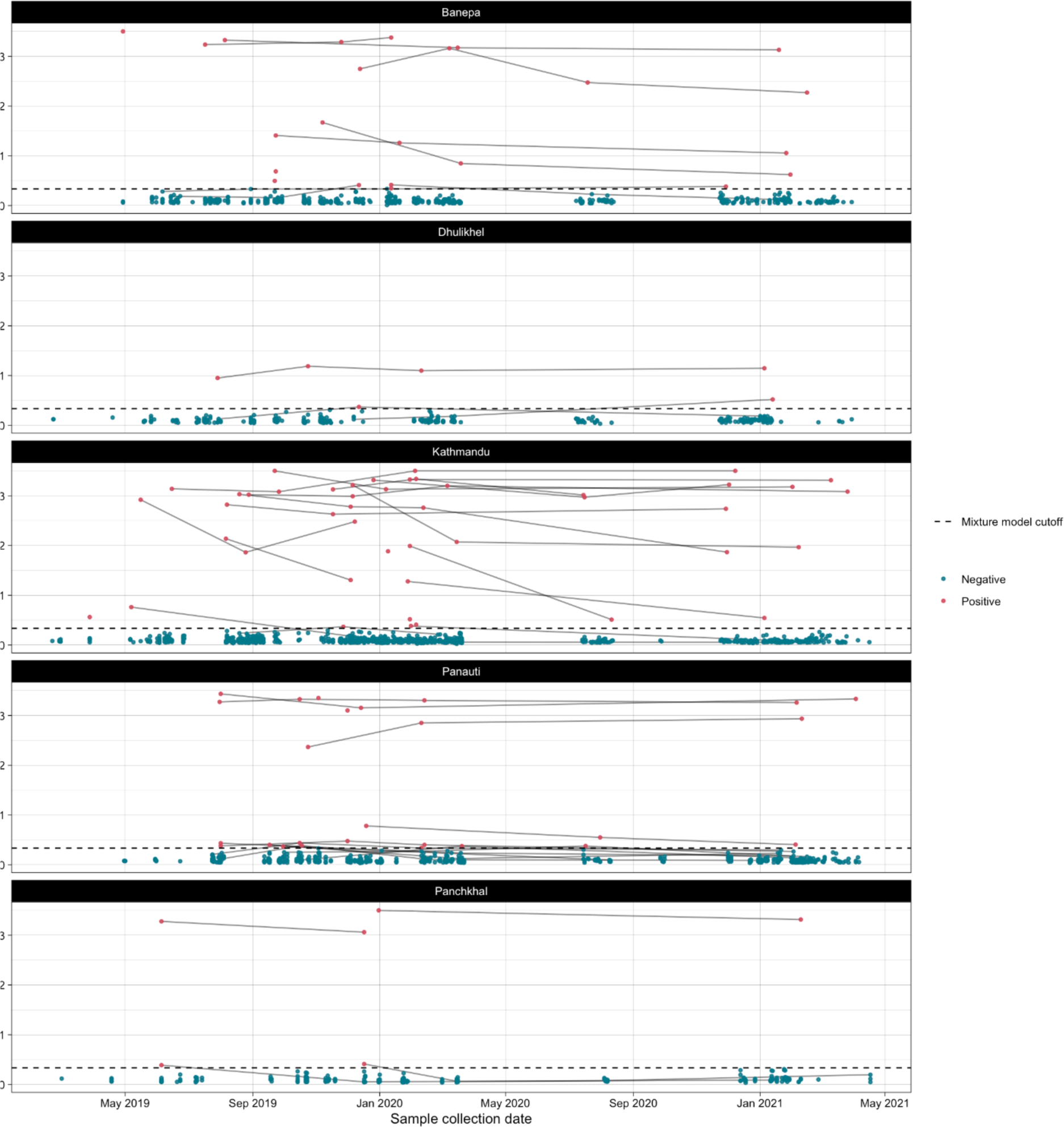
Quantitative antibody responses by date and study site location

**Table 2:** HEV Seroprevalence at baseline visit.

| levels | N() | N seropositive | Crude seroprevalence | Modeled seroprevalence (95% CI) | p value |
| --- | --- | --- | --- | --- | --- |
| Overall | 923 | 44 | 4.8% | • | • |
| Age, categorical |  |  |  |  |  |
| 0-<5 | 199 | 2 | 1.0% | 1.0% (0.3-3.9) | • |
| 5-<10 | 204 | 2 | 1.0% | 1.0% (0.2-3.8) | 0.980 |
| 10-<15 | 223 | 7 | 3.1% | 3.1% (1.5-6.4) | 0.151 |
| 15-25 | 297 | 33 | 11.1% | 11.1% (8.0-15.2) | 0.001 |
| City/town* |  |  |  |  |  |
| Banepa | 167 | 10 | 6.0% | 3.5% (1.7-7.3) | Ref |
| Dhulikhel | 85 | 1 | 1.2% | 0.7% (0.1-5.3) | 0.139 |
| Kathmandu | 401 | 19 | 4.7% | 2.3% (1.2-4.1) | 0.279 |
| Panauti | 197 | 10 | 5.1% | 2.7% (1.3-5.7) | 0.583 |
| Panchkhal | 73 | 4 | 5.5% | 3.6% (1.3-9.9) | 0.973 |
| Gender* |  |  |  |  |  |
| Female | 431 | 19 | 4.4% | 2.4% (1.3-4.3) | Ref |
| Male | 482 | 24 | 5.0% | 2.7% (1.5-4.6) | 0.725 |
| Household monthly income, Nepalese rupees* |  |  |  |  |  |
| <30000 | 412 | 10 | 2.4% | 1.4% (0.7-2.9) | Ref |
| >30000 | 332 | 26 | 7.8% | 4.3% (2.5-7.4) | 0.003 |
| Primary water source* |  |  |  |  |  |
| Municipal | 326 | 11 | 3.4% | 1.9% (0.9-3.9) | Ref |
| Bottled | 246 | 17 | 6.9% | 3.6% (1.9-6.6) | 0.119 |
| Ground | 129 | 5 | 3.9% | 2.3% (0.9-5.8) | 0.760 |
| Private | 40 | 1 | 2.5% | 0.8% (0.1-6.3) | 0.432 |
| Surface | 171 | 9 | 5.3% | 2.9% (1.3-6.3) | 0.368 |
| Household treats drinking water* |  |  |  |  |  |
| No | 472 | 18 | 3.8% | 2.1% (1.2-3.8) | Ref |
| Yes | 440 | 25 | 5.7% | 3.0% (1.7-5.1) | 0.286 |

Note: \*Mixed effect models adjusted for age with a random effect for city/town

**Table 3:** HEV Seroincidence.

|  | Incident seroconversions | Person-years | Seroincidence rate per 100,000 person-years |  |  |
| --- | --- | --- | --- | --- | --- |
|  |  |  | Seroconversions/person-time | Modeled seroincidence* | p-value* |
| Overall | 9 | 822.8 | 10.9 (5.0-20.8) | 10.3 (5.3-19.7) | • |
| Age, categorical |  |  |  |  |  |
| 0-<5 | 0 | 115.2 | 0.0 (0.0-32.0) | 0.00 (0.00-0.31) | • |
| 5-<10 | 2 | 222.7 | 9.0 (1.1-32.4) | 8.73 (2.19-34.89) | Ref |
| 10-<15 | 3 | 214.7 | 14.0 (2.9-40.8) | 13.46 (4.34-41.71) | 0.019 |
| 15-25 | 4 | 270.2 | 14.8 (4.0-37.9) | 13.08 (4.91-34.84) | 0.019 |
| City/town* |  |  |  |  |  |
| Banepa | 2 | 170.5 | 11.7 (1.4-42.4) | 10.5 (2.6-42.6) | Ref |
| Dhulikhel | 2 | 106.9 | 18.7 (2.3-67.6) | 17.1 (4.2-69.2) | 0.625 |
| Kathmandu | 1 | 247.6 | 4.0 (0.1-22.5) | 3.4 (0.5-24.5) | 0.355 |
| Panauti | 4 | 208.9 | 19.1 (5.2-49.0) | 16.8 (6.1-46.3) | 0.588 |
| Panchkhal | 0 | 89.0 | 0.0 (0.0-41.4) | • | 1.000 |
| Gender* |  |  |  |  |  |
| Female | 4 | 389.8 | 10.3 (2.8-26.3) | 8.9 (3.2-24.7) | Ref |
| Male | 5 | 429.4 | 11.6 (3.8-27.2) | 10.3 (4.2-25.5) | 0.831 |
| Household monthly income, Nepalese rupees* |  |  |  |  |  |
| <30000 | 3 | 381.6 | 7.9 (1.6-23.0) | 7.5 (2.4-23.3) | Ref |
| >30000 | 3 | 283.5 | 10.6 (2.2-30.9) | 9.4 (3.0-30.0) | 0.779 |
| Primary water source* |  |  |  |  |  |
| Municipal | 2 | 320.7 | 6.2 (0.8-22.5) | 5.6 (1.4-22.8) | Ref |
| Bottled | 1 | 157.7 | 6.3 (0.2-35.3) | 5.3 (0.7-38.2) | 0.958 |
| Ground | 1 | 138.0 | 7.2 (0.2-40.4) | 6.5 (0.9-46.8) | 0.903 |
| Private | 0 | 26.1 | 0.0 (0.0-141.3) | • | 1.000 |
| Surface | 5 | 176.7 | 28.3 (9.2-66.0) | 24.8 (10.0-61.5) | 0.076 |
| Household treats drinking water* |  |  |  |  |  |
| No | 5 | 442.4 | 11.3 (3.7-26.4) | 10.1 (4.1-24.9) | Ref |
| Yes | 4 | 376.8 | 10.6 (2.9-27.2) | 9.1 (3.3-25.3) | 0.879 |

Note: \*Mixed effect poisson model adjusted for age and repeated measures

**Figure 2:**
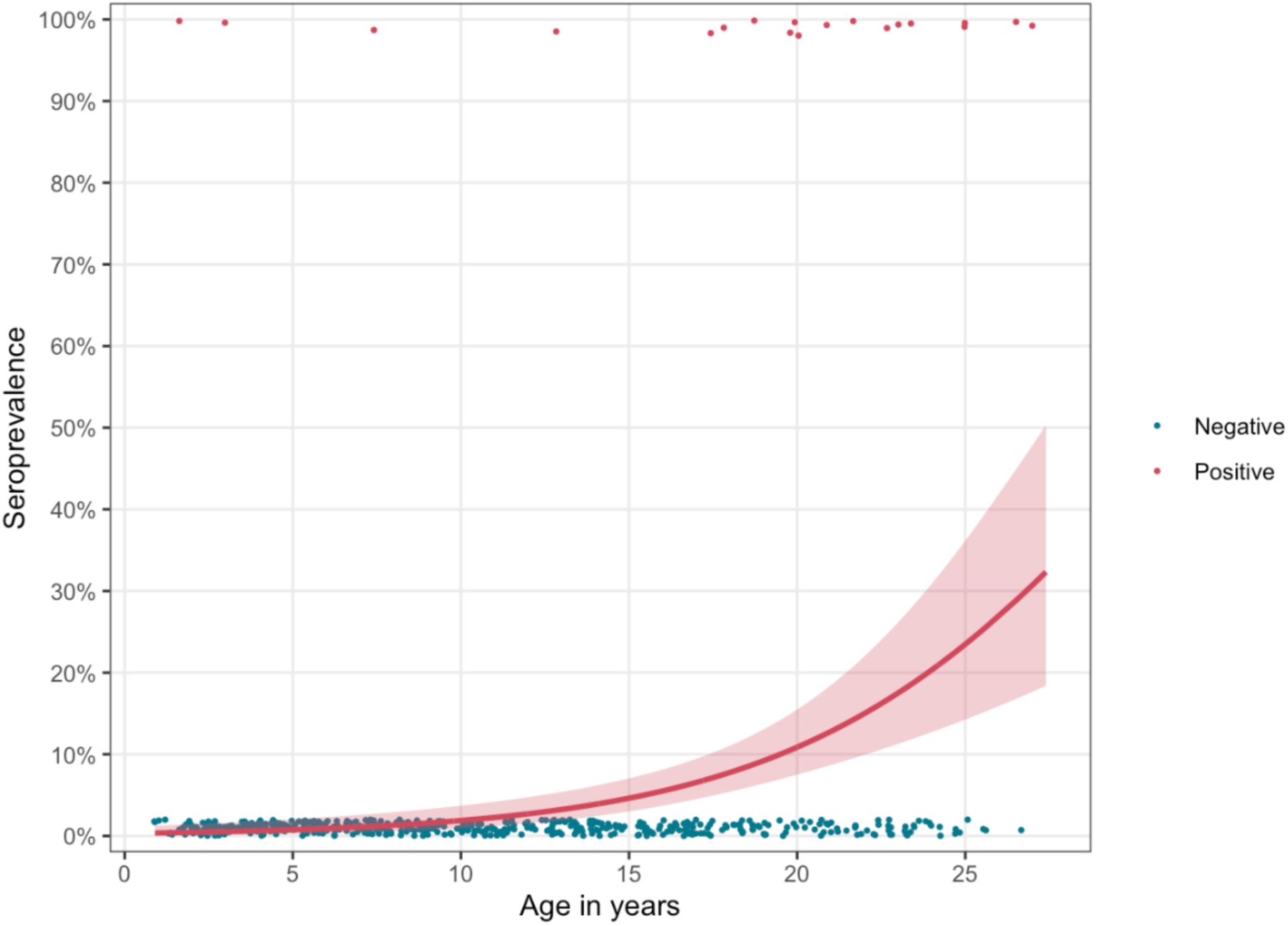
Age-dependent Seroprevalence

**Figure 3:**
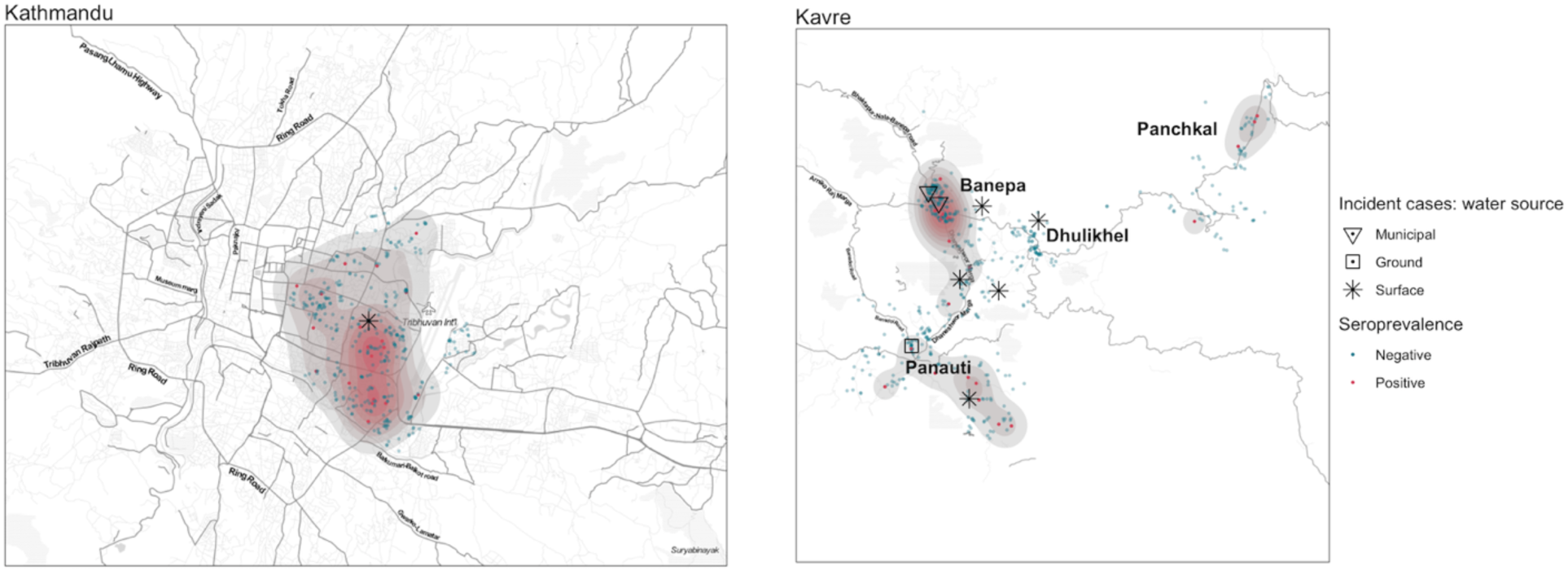
HEV Seroprevalence at baseline and prospective seroconversions across the enrollment areas (Banepa, Panauti, Dhulikhel, Panchkhal: Kavre and Kathmandu) and over the enrollment period of Feb 2019 to Apr 2021

Individuals residing in households with a monthly income exceeding 30,000 Nepalese rupees had a HEV seroprevalence of 4.3% (95% CI 2.5 - 7.5) compared to 1.4% (95% CI 0.7-3.0) for those in households earning below 30,000 Nepalese rupees (p = 0.003). The age-adjusted seroincidence rate for the higher income group was 9.4 (95% CI 3.0 - 30.0) per 1000 person years, and 7.5 (95% CI 2.4-23.3) for the lower income group.

HEV seroprevalence estimates did not vary markedly according to drinking water source, with a seroprevalence of 2.9% (95% CI 1.3-6.3) among individuals whose primary drinking water source was surface water, 3.6% (95% CI 1.9 - 6.6) for bottled water, 1.9% (95%CI: 0.9-3.9) for municipal water, 2.3% (95%CI: 0.9-5.8) for ground water and 0.8% (95%CI: 0.1-6.3) for a private water company. However, the HEV seroincidence rate among individuals drinking surface water was higher than the other groups at 24.8 (95%CI: 9.9 – 61.5) per 1000 person-years compared to 5.6 (95% CI: 1.4-22.8) for municipal water (p = 0.076). Of the 9 incident seroconversions, 5 reported drinking surface water and of these 3 did not treat, 1 treated sometimes (<50% of the time) and 1 reported always treated water by boiling. However, when aggregated there were no differences in HEV seroprevalence or seroincidence according to whether the household treated their drinking water or not. Detailed seroprevalence and seroincidence are provided in Tables 2 and 3.

HEV seroprevalence and seroincidence were not geographically uniform. The seropositive cases are geospatially represented in Figure 2, revealing potential clusters in both Kavre and Kathmandu districts. In Kavre district, seroprevalence was highest in Banepa 3.5% (95% CI 95% CI: 1.7-7.3) and Panchkal 3.6% (95% CI 1.3-9.9). Within Kathmandu, seropositive cases were centered in south-west of Tribhuvan airport, the one incidence seroconversion was also in this area. In Kavre, seroconversions were centered near Banepa (Figure 2).

## DISCUSSION

This representative longitudinal serosurvey reveals ongoing HEV exposure in the Kathmandu Valley of Nepal. With a total of 2513 dried blood samples collected from 923 children and young adults, we observed a seroprevalence of 4.8% and a seroincidence rate of 10.9 seroconversions per 1000 person-years. These findings are consistent with the growing body of evidence suggesting that HEV is endemic in many parts of Nepal (10,11,13,14,20).

Unlike many enteric infections where the youngest are at the highest risk, our study found the highest seroincidence among adolescents and young adults. Among those aged 15-25, 1 in 10 participants had evidence of HEV IgG exposure. This age group is particularly significant as it encompasses women of childbearing age, where the risks of HEV complications and mortality peak (3). This aligns with findings from other studies in Bangladesh (6) and Nepal (20), which reported the highest HEV seroprevalence among oldest enrolled subjects (70-74 in Bangladesh and 30-39 in Nepal). However, with seroprevalence, these other studies were limited in their ability to disentangle age and cohort effects. It’s possible that seroprevalence among older ages reflected higher periods of exposure in the past. In this study, by measuring incident seroconversions, we were able to characterize a genuine higher risk of exposure with age.

The clustering of seropositive cases, especially around Banepa in Kavre and the southwestern region of Tribhuvan airport in Kathmandu, suggests localized outbreaks or common sources of exposure. The majority of incidence seroconversions in Kavre were within five kilometers of Banepa – where a high seroprevalence was observed at baseline. This may suggest propagative transmission from a potential previous outbreak. Such clustering has been observed in other studies (21, 22), and underscores the importance of targeted public health interventions in these regions.

Water source emerged as a significant factor influencing HEV seroincidence. Individuals consuming surface water exhibited a notably higher seroincidence rate compared to those relying on other water sources. This finding aligns with previous studies that have identified contaminated water as a primary transmission route for HEV (21,22). The fact that a majority of the incident seroconversions were observed in individuals who consumed untreated or occasionally treated surface water further underscores the importance of water quality in HEV exposure The spread of HEV to the environment might pollute surface waters, which could act as the source of infection for both humans and animals (23). Moreover, the occurrence of HEV in different water environments, even in industrialized countries with sanitation and safe water supplies, has been documented, emphasizing the need for continuous monitoring and intervention (23).

This study stands out as one of the few representative longitudinal serosurveys specifically designed to capture incidence HEV seroconversions alongside HEV seroprevalence. The variability in seroprevalence estimates across studies, even within identical regions, is well-documented (24). Some of these discrepancies can be attributed to methodologies that overlook the age-dependent nature of seroprevalence and/or ignore antibody decay. Ignoring the waning of antibodies can lead to underestimations of both seroprevalence and seroincidence, contingent on the rate and pattern of antibody decay (25). In our study, the seroincidence derived from the age-dependent cross-sectional visit underestimated the true seroincidence by half. Moreover, seroincidence measures are particularly valuable in identifying recent exposures that could amplify the risk of infection and in pinpointing outbreaks. As an illustration, in our study while there were only minor differences in seroprevalence based on water source, the seroincidence of HEV among individuals consuming surface water was more than quadruple that of those drinking municipal water.

Several limitations are worth noting. Firstly, our cohort did not include individuals above 25 years of age, restricting our ability to comment on the HEV burden in older age groups. However, prior research has indicated that the most significant HEV burden is among young adults(6,20). Our use of dried blood samples, though logistically advantageous, might have increased the limit of detection our serological assays. A recent study comparing HEV IgG responses in dried blood spots to venous blood reported a sensitivity of 81% and a specificity of 97% (26). Given this reduced sensitivity, our findings may underestimate the true HEV seroprevalence and seroincidence by up to 20%. Additionally, our household sampling strategy could have inadvertently omitted migrant populations and residents of informal settlements. Such populations, often faced with subpar water, sanitation, and hygiene conditions, could potentially have a higher HEV seropositivity rate, potentially further biasing our findings towards the null.

In summary, our study underscores the endemic nature of HEV in Nepal, revealing significant variations in seroprevalence and seroincidence based on age, water source, and geography. These insights emphasize the urgency for targeted public health strategies, especially in identified clusters, and spotlight the critical role of safe water practices in curtailing HEV incidence.

## Supporting information

Supplement Figure 1: ELISA optical density (OD) response (represented in log) among baseline samples with cutoff

## Data Availability

All data produced in the present work are contained in the manuscript.

## Acknowledgments

We gratefully acknowledge the study participants for their valuable time and interest in participating in the studies.

