## Supplementary figures and images for "Hepatitis E in Kathmandu Valley: Insights from a Representative Longitudinal Serosurvey"

### Supplement Figure 1: ELISA optical density (OD) response (represented in log) among baseline samples with cutoff

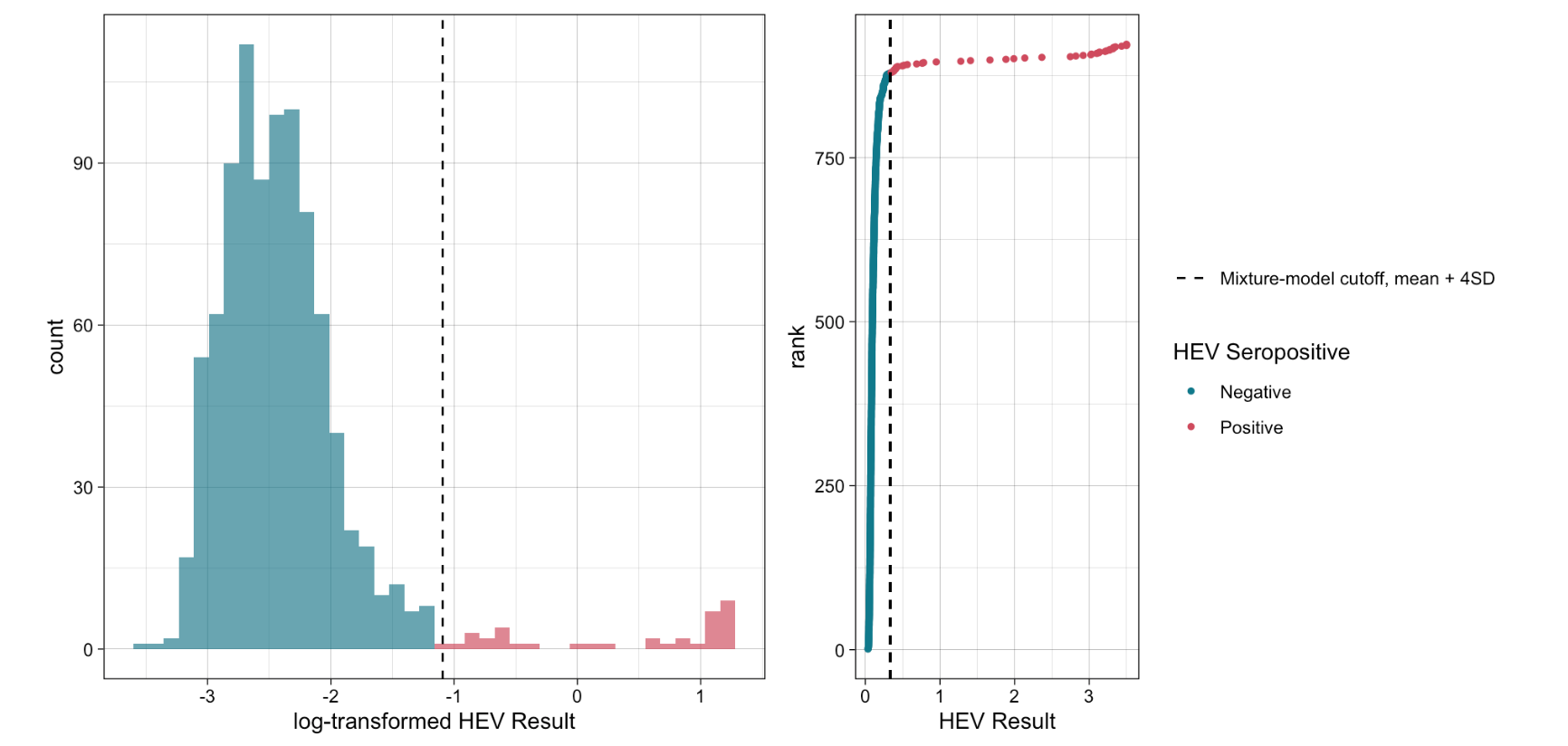
